# Breastfeeding education, early skin-to-skin contact, and other strong determinants of exclusive breastfeeding in an urban population: results from a prospective study

**DOI:** 10.1101/2020.06.12.20129601

**Authors:** Iván Dueñas-Espín, Ángela León-Cáceres, Angelica Álava, Juan Ayala, Karina Figueroa, Vanesa Loor, Wilmer Loor, Mónica Menéndez, David Menéndez, Eddy Moreira, René Segovia, Johanna Vinces

## Abstract

**Objective:** The current study aims to demonstrate independent associations between social, educational, and health practice interventions as determinants of exclusive breastfeeding in an urban Ecuadorian population.

**Design:** Prospective survival analyses.

**Setting:** Ecuadorian mother-child dyads in urban settings.

**Participants:** We followed-up 363 mother-baby dyads who were attended in health care centres in Portoviejo, province of Manabi, for up to 150 days.

**Main outcome measures:** We performed a survival analysis, by setting the time-to-abandonment of exclusive breastfeeding measured in days of life, periodically assessed by phone, as the primary outcome. Crude and adjusted mixed-effects Cox proportional hazards model were performed to estimate hazard ratios (HR) for each explanatory variable.

**Results:** The median time of follow-up (P25 to P75) was 125 (121 to 130) days, and the incidence rate of abandonment of breastfeeding was 8.9 per 1000 persons-days in the whole sample. The three more significant protective determinants of exclusive breastfeeding were *(i)* sessions of prenatal breastfeeding education, with a reduced risk of abandoning the practice of exclusive breastfeeding of 30% (95%CI: 50% to 10%) per each extra session, *(ii)* self-perception of milk production, with a reduced risk of abandoning the practice of exclusive breastfeeding of 57% (95%CI: 50% to 10%) per each increase in the perceived quantity of milk production; and *(iii)* receiving early skin-to-skin contact represented a 90% risk reduction of interrupting breastfeeding (95%CI: 94% to 70%) vs. not receiving.

**Conclusions:** Prenatal education on breastfeeding, self-perception of milk production, and early skin-to-skin contact appears to be strong determinants of exclusive breastfeeding in urban Ecuadorian mother-baby dyads; further, educational level of the mother, excreta management conditions are important determinants too.

**Article summary:** *Strengths and limitations of the study:* One of the most important strengths is that the study, as far as we know, is the first to address the topic in this specific region of the country. Also, this design allows the identification of patterns and elements; helping determine risk factors over time and cause and effect relationships. As we collected data in different intervals, we aimed to minimize recall bias and identify different changes at individual and group level. Our estimates are calculated by proper adjustment of potential confounders, reducing potential sources of confounding bias. This ensured a clear focus and increased validity. This study focused on urban populations as considering that there is a lack of research directed towards this group. However, one limitation can be the lack of representativeness of rural population, in which determinants would be different.^32^ As our research analysed hospital data, another limitation is related to the population in the area that did not go or decided to not deliver in a health centre; considering accessibility and use of the health services as relevant factors. Further research is needed. Also, other potential confounders were not considered, like nutritional factors or blood tests results. However, to our knowledge, there is no reason to consider that such variables could bias the estimates. - This is a longitudinal prospective study developed under real conditions, in an urban Ecuadorian population, in the Coast region, Manabí province.
- A total number of 363 dyads were followed up for 150 days, having collected data at three separated times in three different phone surveys.
- Log-rank tests for equality of survivor functions for assessing differences of actual time in days to abandonment of exclusive breastfeeding were performed.
- A Cox proportional hazards models directed to evaluate the independent association between each explanatory variable and actual time in days to abandonment of exclusive breastfeeding was built, including the estimation of crude and adjusted hazard ratios per each explanatory outcome.
- Several secondary analyses to assess the sensitivity of our estimates with our assumptions regarding biases were conducted, as well as to test for model misspecifications.

## Introduction

Breastfeeding promotion is a highly cost-effective health intervention with individual, social, and economic advantages.^1^ Benefits from exclusive breastfeeding have been extensively demonstrated during childhood and, even, adulthood.^2–4^ Mothers who breastfeed have better health outcomes, like a decreased risk of breast and ovarian cancer, and of hip fractures, and osteoporosis in the postmenopausal period.^5^

Several social and economic determinants of breastfeeding have been demonstrated;^6^ among these, socioeconomic barriers preclude a proper breastfeeding for infants and children.^6^ Specifically, Ecuador has several difficulties in implementing policies towards exclusive six months breastfeeding.^7,8^ There are difficulties in the access to services which promote breastfeeding, such as: lactation support rooms, milk banks, exposure to edu-communicational interventions for breastfeeding promotion, among others. Furtherly, industry has significant influence on breastfeeding decisions, as it was showed that 18% of new Ecuadorian mothers participate in industry-sponsored social groups and activities.^9^

As a result, Ecuador has a low rate of exclusive six months breastfeeding according to two massive national surveys; specifically, in the province of Manabí, the *montubio* ethnic group presents only 23% of children reaching up to six months of exclusive breastfeeding.^8^ Despite several policies and regulations for health care services directed to improve such indicator exist, any of them has been effectively applied; thus, this enhances the need for applying more efforts, specially, at the health care area.^10^ In this province, 69% of the population define themselves as *mestizos*, while the second majority (19%) are *montubios*.^11^ Historically, the latter is considered a united ethnic group with a common background of oppression and a shared need for recognition of rights and targeted governmental interventions.^12^ Additionally, in this area, there is a 10.2% of illiteracy, majority of the population has no social insurance, and women are less economically active than men.^11^

In the prenatal, natal, and postnatal health care areas; several determinants heavily influence the maintenance of breastfeeding. In that sense, the United Nations Children’s Fund (UNICEF) and the World Health Organization (WHO) launched the Baby-friendly hospital initiative (BFHI) – a strategy at the health care level for enhancing good practices towards improved adherence to breastfeeding– which is well recognized as a cost-effective way towards the promotion and protection of breastfeeding.^13^ BFHI has been aimed to: *(i)* improve the quality and comprehensiveness of prenatal care, *(ii)* promote humanized delivery and adequate newborn care, *(iii)* improve the quality of care for obstetric and neonatal emergencies, *(iv)* prevent vertical transmission of HIV and syphilis, and *(v)* promote, support, and protect breastfeeding.

Specifically, the early skin-to-skin contact strategy has been demonstrated as effective for improving exclusive and nonexclusive breastfeeding rates,^14^ considering it as a strong determinant of breastfeeding. Nevertheless, to our knowledge, this practice has not been tested/proved as a determinant of breastfeeding in Ecuador. Consequently, the current study aims to demonstrate independent associations between sociodemographic characteristics, educational background, and health practice interventions as determinants of exclusive breastfeeding in an urban Ecuadorian population.

## Methods

### Design

Prospective survival analyses.

### Population and sample

We initially recruited 400 mother-baby dyads who were attended in either a) one hospital or b) six primary health care facilities in Portoviejo, in the province of Manabí. Given that the determinants of breastfeeding are different for premature infants, we excluded from the follow-up those dyads in which the infant was born at <37 weeks of gestational age. A total number of 363 dyads were followed up for 150 days (see the study flowchart in the **Figure 1S** of the **online supplementary material**).

We included in the study dyads in which: *(i)* mothers were at immediate or mediate puerperium, it means from delivery to <40 days postpartum and who were on exclusive breastfeeding; *(ii)* whose neonates were alive; *(iii)* mothers who can read and did not have physical, motor, intellectual or visual disabilities; *(iv)* mothers who have not had or were not contraindicated to carry out breastfeeding (e.g. HIV, active infections of the mammary gland, active pulmonary TB). We excluded dyads in which: *(i)* neonates died; *(ii)* mothers who cannot read and/or had physical, motor, intellectual or visual disabilities; *(iii)* mothers who have had contraindication to carry out breastfeeding; and *(iv)* mothers who have not signed the informed consent and/or who did not want to participate in the study.

### Main outcome

We prospectively followed-up the dyads, and performed a survival analysis, by setting the time-to-abandonment of exclusive breastfeeding measured in days of life. We periodically assessed, by phone, the date in which mothers told that other food or liquid than breast milk was given to the baby, according to the WHO’s definition of exclusive breastfeeding.^15^

### Measurements

Socio demographic data and health practice interventions were collected through telephone survey. Data collection was done at three times, at the first contact, at the second and at the fourth month, in three different phone surveys.

Several variables were obtained: mother’s age, marital status, education, working status, type of health insurance, and other socioeconomic conditions, prenatal care variables (number of prenatal care office visits, sessions of education about breastfeeding, obstetric risk), natal care like type of delivery, health care practice during delivery (skin-to-skin contact, joint accommodation, timely ligation of the umbilical cord, breastfeeding within the first hour of life); and, infant variables, (sex, gestational age, birth weight in grammes, self-perception of milk production).

### Statistical analyses and sample considerations

Using the early skin-to-skin contact variable as the main explanatory variable, we performed a sample calculation, a posteriori, by using next parameters: considering that the incidence rate of abandonment of exclusive breastfeeding was 2.8 events per 1000 patient-days, and with an α (two-tailed) = 0.05 and a β = 0.2, we found that the minimal patients in a group with skin-to-skin contact (exposed) should be of at least 204 patients, and in the unexposed, of 42 patients (ref); being the final sample constituted by 363, with 302 exposed patients, and 61 unexposed patients, assuring statistical power.

Descriptive statistics were performed using percentages for categorical variables and median and P25 to P75 for discrete variables. We performed log-rank tests for equality of survivor functions for assessing differences of actual time in days to abandonment of exclusive breastfeeding. Then, we estimated crude and adjusted hazard ratios (HR) per each explanatory outcome. In that sense, we built multivariate Cox proportional hazards models to evaluate the independent association between each explanatory variable and actual time in days to abandonment of exclusive breastfeeding. We built a saturated model including all the individual covariates. Then, based on the researchers’ criteria, we eliminated covariates from significant covariates that were retained in the model. HR were estimated. Confidence intervals (95%CI) of the HR and their corresponding p-values were calculated. Once the parsimonious model was obtained, we compared both models and chose the “final” model, according to its level of significance from the likelihood ratio test. To assess the effects from the socioeconomic level we estimated HR by mixed effects from the Cox proportional hazards model. Given the small number of missing data (there were missing values in <1% of the whole database), we employed complete case analysis in estimating statistical associations.

To test for potential effect modification, we stratified main analysis according to the sex of the infant. Also, we performed several secondary analyses to assess the sensitivity of our estimates with our assumptions regarding biases, as well as to test for model misspecifications. First, we stratified by sex of the infant; second, we ran the final model excluding: *(i)* dyads with single mothers, *(ii)* those without schooling or only basic education, *(iii)* those dyads from high socioeconomic level, and *(iv)* those dyads in which the infant was born by a C-section.

This study was conducted with the approval of the Research Ethics Committee in Humans (RECH) of the Pontificia Universidad Católica del Ecuador (code number: 2018-48-EO).

## Results

From the women that took part in the survey, the average age was 23 years old; from which a majority declared having a couple (73%). Around 61% of them reported having middle education, belonging to a medium-high socioeconomic status (48%), and almost all of them were unemployed with no health insurance (82%). Regarding the type of delivery, there is a high percentage of women who had a C-section (49%), reporting also high obstetric risk (34%). Relevantly, when asked about health care practices during delivery, results show that there are high percentages of women that received joint accommodation (93%), skin to skin contact (83%), and timely ligation of the umbilical cord (93%). Nevertheless, only 63% of women breastfed within the first hour of life though majority reported perceiving enough quantity of milk production for their babies (37%), see **Table 1**.

**Table 1.** Baseline characteristics.

| Baseline characteristics<sup>a</sup> | Patients<br>n= 363 |
| --- | --- |
| **Mother's age (years), P50 (P25-P75)** | 23 (19 to 28) |
| **Mother's marital status** |  |
| Single, n (%) | 24 (7) |
| With couple, n (%) | 264 (73) |
| Separated, n (%) | 10 (3) |
| Married, n (%) | 60 (17) |
| Divorced, n (%) | 4 (1) |
| Widow, n (%) | 1 (<1) |
| **Education** |  |
| Without schooling, n (%) | 1 (<1) |
| Basic education, n (%) | 74 (20) |
| Middle education, n (%) | 223 (61) |
| Higher education, n (%) | 65 (18) |
| **Working status** |  |
| Full occupation, n (%) | 33 |
| Unemployment, n (%) | 309 |
| Underemployment, n (%) | 21 |
| **Health insurance** |  |
| Social Security, n (%) | 53 (14) |
| Other than social security, n (%) | 13 (4) |
| None, n (%) | 297 (82) |
| **Socioeconomic conditions** |  |
| Elimination of excreta by toilet, n (%) | 337 (93) |
| Disposal of excreta by latrine, n (%) | 26 (7) |
| **Socioeconomic level<sup>b</sup>** |  |
| High level, n (%) | 70 (19) |
| Medium high, n (%) | 175 (48) |
| Medium, n (%) | 97 (27) |
| Medium low, n (%) | 21 (6) |
| **Number of prenatal care office visits, P50 (P25 to P75)** | 2 (1 to 3) |
| **Sessions of education about breastfeeding, P50 (P25 to P75)** |  |
| **Obstetric risk<sup>c</sup>** |  |
| No risk, n (%) | 80 (22) |
| Low risk, n (%) | 119 (33) |
| High risk, n (%) | 125 (34) |
| Very high risk, n (%) | 39 (11) |
| **Type of delivery** |  |
| Eutocic vaginal delivery, n (%) | 180 (50) |
| Dystocic vaginal delivery, n (%) | 4 (1) |
| Elective C-section, n (%) | 122 (34) |
| Emergent C-section, n (%) | 56 (15) |
| **Health care practice during delivery** |  |
| Skin-to-skin contact, n (%) | 302 (83) |
| Joint accommodation, n (%) | 336 (93) |
| Timely ligation of the umbilical cord, n (%) | 339 (93) |
| Breastfeeding within the first hour of life, n (%) | 245 (67) |
| **Infant variables** |  |
| Male sex, n (%) | 202 (56) |
| Gestational age, P50 (P25 to P75) |  |
| Birth weight in g, mean (SD) | 3128 (383) |
| **Self-perception of milk production** |  |
| Very little quantity, n (%) | 6 (2) |
| Little quantity, n (%) | 50 (14) |
| Moderate quantity, n (%) | 72 (20) |
| Enough quantity, n (%) | 136 (37) |
| More than enough quantity, n (%) | 99 (27) |

m=mean; SD=standard deviation. <sup>a</sup>The categories used (man = male gender) are the same as those collected by the National Institute of Statistics and Censuses. There were missing values in <1% of the whole database. <sup>b</sup>The Graffar-Méndez Scale was applied; it uses the mother's level of instruction, source of family income and housing conditions. <sup>c</sup>Obstetric risk was categorized according to the number of health risk conditions during pregnancy (i.e. no risk = any condition, low risk = 1 condition, high risk = 2 conditions, and very high risk = 3 or more conditions).

The median time of follow-up (P25 to P75) was 125 (121 to 130) days, and the incidence rate of abandonment of breastfeeding was 8.9 per 1000 persons-days in the whole sample. When measuring the adjusted association between several factors and the maintenance of breastfeeding, results showed a strong association (p-value < 0.01) between mothers’ education and the interruption of breastfeeding. Thus, mothers with higher education have 2.6 times higher risk [95% confidence interval (95%CI): 1.2 to 5.9] of abandoning breastfeeding. Mothers within this category of education represented the 18% of the total sample. While considering the socioeconomic conditions and relating these to access to water and sanitation services, 93% of women were eliminating excreta by toilet; nevertheless, the rest (7%) who used a latrine, were 1.8 times more likely to abandon breastfeeding, but this association was not statistically significant (95%CI: 0.7 to 5.2), see **Table 2**.

**Table 2.** Crude and adjusted hazard ratios, estimated by mixed-effects Cox Proportional Hazards Models per each explanatory variable.

| Explanatory variables | Crude hazard ratios (95% CI) | p-value | Adjusted hazard ratios (95% CI) |  |  |  |
| --- | --- | --- | --- | --- | --- | --- |
|  |  |  | Saturated model | p-value | Parsimonious model | p-value |
| **Mother's age (per each year of increasing)** | 0.9 (0.9 to 1.0) | 0.69 | 1.0 (0.9 to 1.1) | 0.85 | - | - |
| **Mother's marital status** |  |  |  |  |  |  |
| Single (other marital status ref.) | 3.0 (1.2 to 7.6) | 0.02 | 1.8 (0.5 to 6.4) | 0.18 | 1.9 (0.6 to 6.1) | 0.30 |
| **Education** |  |  |  |  |  |  |
| Higher education, (lower than higher is the ref.) | 1.9 (0.9 to 4.0) | 0.07 | 3.0 (1.3 to 7.1) | 0.01 | 2.6 (1.2 to 5.9) | 0.01 |
| **Working status** |  |  |  |  |  |  |
| Underemployment (otherwise is the ref.) | 1.0 (0.3 to 3.2) | 0.97 | 1.1 (0.3 to 4.0) | 0.18 | - | - |
| **Socioeconomic conditions** |  |  |  |  |  |  |
| Disposal of excreta by latrine (otherwise is the ref.) | 3.5 (1.5 to 7.9) | <0.01 | 2.0 (0.7 to 5.9) | 0.19 | 1.8 (0.7 to 5.2) | 0.21 |
| **Sessions of breastfeeding education (per each extra session)** | 0.8 (0.7 to 1.1) | 0.11 | 0.7 (0.5 to 0.9) | 0.01 | 0.7 (0.5 to 0.9) | 0.01 |
| **Obstetric risk** |  |  |  |  |  |  |
| Risk score (per each extra risk) | 1.2 (0.8 to 1.7) | 0.35 | 1.2 (0.8 to 1.7) | 0.48 | - | - |
| **Health care practice during delivery<sup>a</sup>** |  |  |  |  |  |  |
| Skin-to-skin contact (otherwise is the ref.) | 0.2 (0.1 to 0.4) | <0.01 | 0.2 (0.1 to 0.3) | <0.01 | 0.1 (0.06 to 0.30) | <0.01 |
| Joint accommodation (otherwise is the ref.) | 0.7 (0.2 to 1.9) | 0.44 | - | - | - | - |
| Timely ligation of the umbilical cord (otherwise is the ref.) | 0.4 (0.2 to 1.1) | 0.09 | - | - | - | - |
| Breastfeeding within the first hour of life (otherwise is the ref.) | 0.6 (0.3 to 1.2) | 0.18 | - | - | - | - |
| **Self-perception of milk production** |  |  |  |  |  |  |
| Very little quantity (ref.) | 1 | - | 1 | - | 1 | - |
| Little quantity | 0.44 (0.15 to 1.35) | 0.15 | 0.73 (0.20 to 2.69) | 0.63 | 0.70 (0.20 to 2.40) | 0.56 |
| Moderate quantity | 0.13 (0.03 to 0.42) | <0.01 | 0.13 (0.03 to 0.51) | <0.01 | 0.12 (0.03 to 0.47) | <0.01 |
| Enough quantity | 0.06 (0.02 to 0.19) | <0.01 | 0.07 (0.02 to 0.31) | <0.01 | 0.08 (0.02 to 0.29) | <0.01 |
| More than enough quantity | 0.04 (0.01 to 0.17) | <0.01 | 0.08 (0.02 to 0.39) | <0.01 | 0.09 (0.02 to 0.40) | <0.01 |
| p-for-trend | 0.41 (0.30 to 0.55) | <0.01 | 0.42 (0.30 to 0.59) | <0.01 | 0.43 (0.31 to 0.59) | <0.01 |
| **Depressive symptoms by PHQ2** |  |  |  |  |  |  |
| Three or more points in the score (<3 is the ref.) | 1.7 (0.2 to 12.5) | 0.59 | 2.6 (0.3 to 24.3) | 0.41 | - | - |
| **Infant variables** |  |  |  |  |  |  |
| Male sex (female is the ref.) | 1.3 (0.7 to 2.6) | 0.38 | 1.2 (0.6 to 2.5) | 0.54 | - | - |
| Gestational age in weeks (per each increase in tertile) | 1.4 (1.0 to 2.2) | 0.07 | 1.5 (0.9 to 2.4) | 0.11 | 1.5 (0.9 to 2.4) | 0.08 |
| First tertile (ref.) | 1 | - | 1 | - | 1 | - |
| Second tertile | 0.9 (0.4 to 2.2) | 0.86 | 1.24 (0.47 to 3.28) | 0.67 | 1.21 (0.47 to 3.12) | 0.69 |
| Third tertile | 1.9 (0.9 to 4.3) | 0.09 | 2.18 (0.85 to 5.60) | 0.10 | 2.26 (0.92 to 5.58) | 0.07 |
| Birth weight, per each g of increase | 0.9 (0.9 to 1.0) | 0.38 | 1.0 (0.9 to 1.0) | 0.70 | - | - |
| Any complication at birth (no complication ref.) | 1.7 (0.5 to 5.4) | 0.39 | 2.1 (0.6 to 8.1) | 0.27 | - | - |

<sup>a</sup>There was found collinearity in the models between early skin-to-skin contact and: joint accommodation, timely ligation of the umbilical cord, and breastfeeding within the first hour of life; those variables were excluded from modelling the saturated model (see supplementary material for details).

In the other hand, our study shows a strong association between protective variables and the time on exclusive breastfeeding practice. The three more significant (p-value < 0.01) were *(i)* sessions of breastfeeding education, with a reduced risk of abandoning the practice of exclusive breastfeeding of 30% (95%CI: 50% to 10%) per each extra session during prenatal care, *(ii)* self-perception of milk production, with a reduced risk of abandoning the practice of exclusive breastfeeding of 57% (95%CI: 50% to 10%) per each increase in the perception of milk production at the mediate puerperium (**Figure 1**); mainly, women that perceived having enough or more than enough quantity of breast milk, are 92% and 91% less likely to desert the breastfeeding practice, respectively (95%CI: 71% to 98%; and, 60% to 98%, respectively); and *(iii)* application of early skin-to-skin contact which represented a 90% risk reduction of interrupting breastfeeding (95%CI: 94% to 70%), see **Figure 2**. Interestingly, adding other comprehensive healthcare practices, different to early skin-to-skin contact, in the models resulted in collinearity; thus, we excluded them from the modelling (**Tables 1S and 2S**).

**Figure 1.**
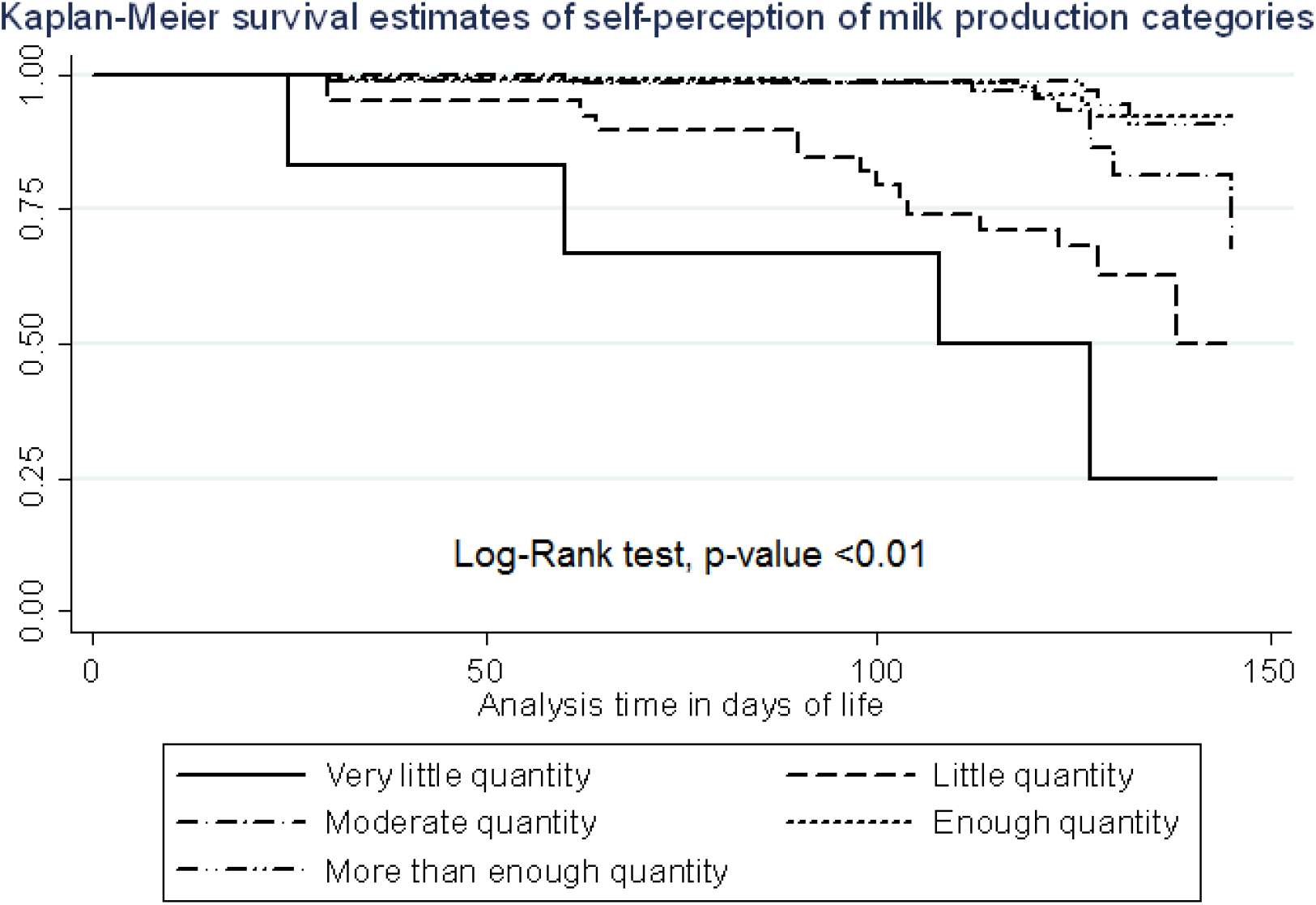
Kaplan-Meier survival estimates of self-perception of milk production categories.

**Figure 2.**
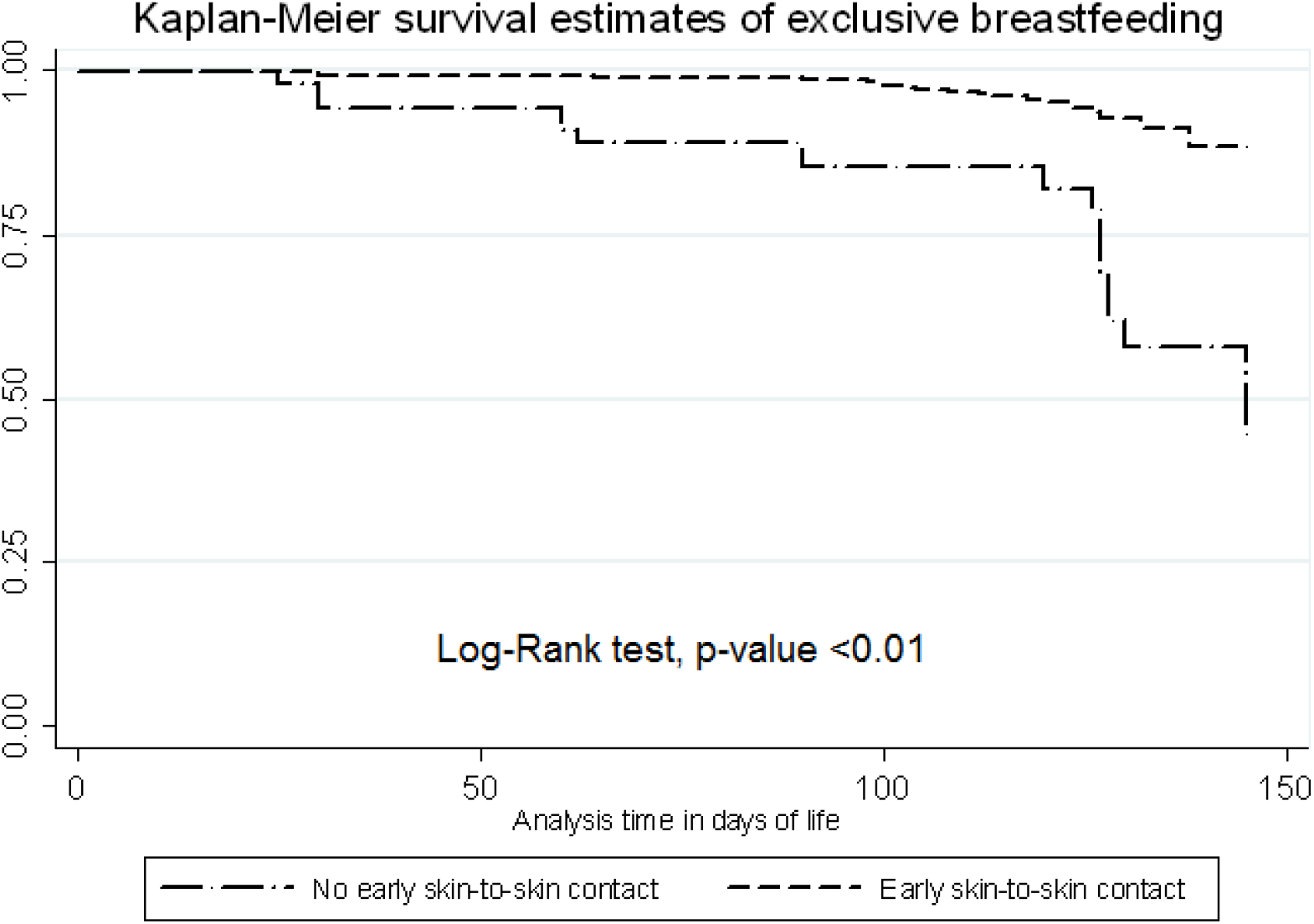
Kaplan Meier survival estimates of exclusive breastfeeding by early skin-to-skin contact categories.

The effect of the determinants on time-to-abandonment did not change after stratifying by sex of the infant (**Table 3S**). Sensitivity analyses yielded very similar results; despite we found a stronger association between single marital status and time-to-abandonment (HR=6.5) when we excluded those dyads in which the infant born by C-section, the confidence interval widened (95%CI: 1.7 to 24.7), probably because there were only 10 single women who gave birth by caesarean delivery (**Table 4S**).

## Discussion

As several other studies have shown, many factors determine the decisions of women to breastfeed their babies.^16–18^ Consequently, the duration of this period and the practices around it also vary. As this is the first study of this type to be conducted in this specific area, the results of our study show that the educational level of the mother, excreta management conditions, self-perception of milk production, prenatal education on breastfeeding, and early skin-to-skin contact appear to be strong determinants of exclusive breastfeeding in an urban population

Early skin to skin contact has been demonstrated as a strong determinant of exclusive breastfeeding by a systematic review.^14^ A prospective cohort study conducted in Poland also contributed to this statement, by indicating that at least 30 minutes of skin-to-skin contact led babies to be 1.2 month longer exclusively breastfed.^19^

Despite some studies^20,21^ evidenced that a low level of education is related to an early abandonment of exclusive breastfeeding, some others, including our study, denote the opposite. In this sense, mothers with less education might tend to introduce other foods earlier, in contrast to mothers who have higher education level. This might be related to increased access of information and recognition of the benefits of exclusive breastfeeding for mothers with higher education. Mothers with higher levels of education and those who had received guidelines on breastfeeding from a health professional show better knowledge and breast feeding practices.^22^ On the other hand, mothers with higher education also present a risk of abandoning breastfeeding; this can be related, among others, to their labour or occupational status in which time and spaces could be influential factors; specifically in urban contexts. This finding was also showed in studies conducted in Ethiopia^23^ and Bangladesh.^24^

Additionally, women’s socioeconomic status might also lead them to purchase breast-milk substitutes and consequently avoid exclusive breastfeeding.^25^ Even if majority of women in our study reported to be unemployed, there can be an association with the informal economy or unpaid housework, in which adequate maternity and workplace entitlements for breastfeeding are non-existent. Thus, interaction with diverse factors might be present during the different analysis and no generalization can be made, as these relations might appear differently according to the context or the individual.

The main reason reported by the mothers for delaying early initiation of breastfeeding (EIBF) was the delay due to the separation of mother and child immediately after birth (55%). Similar findings have been reported by previous studies.^26–28^ This can be related to the number of women that had a C-Section (49%) which can be associated to a prolonged separation of the dyad. Specifically, if this part of the population reported having obstetric risk, entailing longer periods of recovery for the mother and thus a possible longer separation. The influence of C-Sections on EIBF was also reported as a main factor in a prospective multi-country cohort study.^29^

As the benefits of breastfeeding can be seen, at short and long-term, mainly on the social, economic, and environmental spheres; national health authorities should work towards the promotion, protection, and support of the practice with a special emphasis on the political advocacy at the multisectoral and intersectoral levels; leveraging financial investment, resource mobilization, and the organization of supportive networks. A key example is the adaptation, implementation, and evaluation of the BFHI. On the other hand, it is crucial to maintain and implement strong policies which restrict marketing of breastmilk substitutes, at the public and private level; as also suggested by relevant scientific evidence.^1^

Health care professionals must improve the adherence and implementation of comprehensive health practices during delivery, especially considering that current evidence demonstrates that the application of early skin-to-skin contact could importantly enhance the initiation and maintenance of breastfeeding. To our knowledge, this is the first study that demonstrate such association in the Ecuadorian context.

In addition, health care professionals should consider education, sociodemographic characteristics, and cultural factors when counselling mothers to breastfeed. Furthermore, including male partners in the educational breastfeeding sessions could enhance adherence to exclusive breastfeeding practices; as indicated in a study conducted in the United States in which partners posed a positive effect on the mother’s attitudes and intentions to breastfeed.^30^ Relevantly, the applicability of educative sessions should be performed on the antenatal and postnatal period, and the healthcare services provided accordingly.

Given that we found collinearity between early skin-to-skin contact with each one of the other comprehensive health practices during delivery, obstetricians and general practitioners should consider assuring that, at least, skin-to-skin contact is applied in every delivery, including cases of C-sections. This strategy should be accompanied by the initiation of breastfeeding within the first hour, mainly but not exclusively, for the cases in which there are low birth weight and premature babies.^31^

This is a longitudinal prospective study, developed under real conditions from an Ecuadorian urban population. Our estimates are calculated by proper adjustment of potential confounders, reducing potential sources of confounding bias. Probably, the most relevant limitation was the lack of representativeness of rural population, in which determinants would be different.^32^ As our research analysed hospital data, another limitation is related to the population in the area that did not go or decided to not deliver in a health centre; considering accessibility and use of the health services as relevant factors. This, especially considering that exposure to antenatal care and breastfeeding education sessions report a positive relation to adherence to breastfeeding in the postnatal period. Also, other potential confounders were not considered, like nutritional factors or blood tests results. However, to our knowledge, there is no reason to consider that such variables could bias the estimates. Finally, Ecuador is a country with four different regions and an immense diversity not only geographically, but socially, economically, culturally, and ethnically. Despite the study shows a reality that might be applicable to some other similar contexts in the Ecuadorian Coast region, results cannot be fully generalized to other places with different socio-economic, cultural, or geographic contexts.

## Conclusion

Prenatal education on breastfeeding, self-perception of milk production, and early skin-to-skin contact appears to be strong protectors of exclusive breastfeeding in urban Ecuadorian mother-baby dyads; further, educational level of the mother and socioeconomic conditions like excreta management are important social determinants too.

It is imperative that policies focus on social inequities to reduce barriers towards breastfeeding. Health care strategies, as the Baby Friendly Hospitals postulates, have a pivotal role for improving breastfeeding maintenance. As the study focused on analysing the independent associations between social, educational, and health practice interventions as determinants of exclusive breastfeeding, further research related to motivational determinants and how cultural beliefs and practices influence the health seeking behaviour of individuals and communities, is needed to complement the full panorama of breastfeeding determinants in this urban context.

In conclusion, to improve adherence to exclusive breastfeeding and enjoy the social and economic benefits not only for the mother and the child but for the society as a whole, supportive policies from the educational, economic, employment, infrastructure, housing, sanitation, and basic services areas are urgently needed. This, considering that the inclusion of health in all policies will direct efforts towards the population which need it the most.

## Footnotes

## Acknowledgments

Authors thank the input from the *Instituto de Salud Pública* from PUCE.

## Contributorship statement

IDE takes responsibility for (is the guarantor of) the content of the manuscript. All authors contributed to *(i)* the conception, design of the study, acquisition of the data, analysis and interpretation of results, *(ii)* critically revised the article, *(iii)* approved the final version to be published, and *(iv)* agreed to be accountable for all aspects of the work.

## Ethical considerations

This study was conducted with the Research Ethics Committee in Human (RECH) of the *Pontificia Universidad Católica del Ecuador* approval (code number: 2018-48-EO).

## Patient consent for publication

yes.

## Data sharing statement

The project is currently making available the main study results and data by making a request through the correspondence author.

## No Patient and Public Involvement

This research was done without patient involvement. Patients were not invited to comment on the study design and were not consulted to develop patient relevant outcomes or interpret the results. Patients were not invited to contribute to the writing or editing of this document for readability or accuracy.

